# Constitutional thinness and anorexia nervosa differ on a genomic level

**DOI:** 10.1101/2021.03.08.21253137

**Authors:** Christopher Hübel, Mohamed Abdulkadir, Moritz Herle, Alish B. Palmos, Ruth J.F. Loos, Gerome Breen, Nadia Micali, Cynthia M. Bulik

## Abstract

Constitutional thinness and anorexia nervosa are both characterised by persistent, extremely low weight with body mass indices (BMI) below 18.5 kg/m^2^. Individuals with anorexia nervosa concurrently show distorted perceptions of their own body and engage in weight-loss behaviours, whereas individuals with constitutional thinness typically wish to gain weight. Both are heritable, share genomics with BMI, but have not been shown to be genetically correlated with each other. We aim to differentiate between constitutional thinness and anorexia nervosa on a genomic level.

First, we estimated genetic correlations between constitutional thinness and eleven psychiatric disorders and compared them with anorexia nervosa using publicly available data. Second, we identified individuals with constitutional thinness in the Avon Longitudinal Study of Parents and Children (ALSPAC) by latent class growth analysis of measured BMI from 10 to 24 years (*n* = 8,505) and assigned polygenic scores for eleven psychiatric disorders and a range of anthropometric traits to evaluate associations.

In contrast to anorexia nervosa, attention deficit hyperactivity disorder (*r*_gAN_ = 0.02 vs. *r*_gCT_ = −0.24) and alcohol dependence (*r*_gAN_ = 0.07 vs. *r*_gCT_ = −0.44) showed a statistically significant negative genetic correlation with constitutional thinness. A higher polygenic score for posttraumatic stress disorder was associated with an increased risk of constitutional thinness in the ALSPAC cohort (OR = 1.27; *Q* = 0.03) whereas posttraumatic stress disorder shows no genetic correlation with anorexia nervosa (*r*_g_ = −0.02). Overall, results suggest that constitutional thinness is different from anorexia nervosa on the genomic level.

## Introduction

Individuals with constitutional thinness have persistently low body mass indices (i.e., BMIs < 18.5 kg/m^2^) [1] as defined by the World Health Organization (WHO) [2] and show no symptoms of disordered eating [3]. Persistence of low BMI is an important component of the phenotype and requires longitudinal assessment. In some cases, the phenotypic definition is augmented by a family history of thinness as an additional criterion [4]. However, definitions of constitutional thinness vary considerably across studies [5]. The prevalence of constitutional thinness is unknown [1], but is likely to be less than 0.4% for men and less than 2.7% for women (underweight from all causes) [6]. Gaining weight in constitutional thinness is complicated as individuals may be resistant to diet supplementation (e.g., with high-fat foods), as seen in a small controlled study [7].

Constitutional thinness shares only a few features with the restricting subtype of anorexia nervosa. A BMI < 18.5 kg/m^2^ is the main shared symptom between these traits, as individuals with anorexia nervosa display restrictive eating behaviours, body dissatisfaction, and distorted perceptions of their own body [8]. Although individuals with constitutional thinness may also display body dissatisfaction, they typically want to gain, rather than lose weight. However, the risk for potential misdiagnosis exists [1].

Small studies on food intake and energy expenditure indicated an equilibrated energy balance in individuals with constitutional thinness, with higher resting metabolic rate (RMR) and total energy expenditure (TEE) than healthy normal weight controls when adjusted for differences in metabolic mass [4]. However, findings are mixed, as similar levels of resting energy expenditure [9] and similar resting metabolic rates (RMR) as in controls have also been reported [10].

Individuals with constitutional thinness have been shown to have lower levels of general psychopathology reporting lower body dissatisfaction and drive for thinness [3], and fewer depressive and anxiety symptoms [11] in comparison to controls. However, constitutional thinness has been reported to be associated with an adverse mental health profile in men, potentially due to the discrepancy between their body size and the masculine muscular ideal [12]. From a social perspective, early studies did not find associations between constitutional thinness and educational attainment, parents’ education, or marital history [3].

In comparison to anorexia nervosa, which can affect multiple organ systems throughout the human body, individuals with constitutional thinness are generally regarded as healthy. Women with constitutional thinness have preserved menses (without contraceptive use) and good reproductive health [1]. However, on average their menarche occurs one year later than in non-thin controls [3]. Endocrine disturbances are rare [13], as individuals with constitutional thinness have comparable stress and sex hormone concentrations as controls [14]. Compared with controls, individuals with constitutional thinness do present lower concentrations of leptin; however, they are higher than those observed in anorexia nervosa. In addition, the circadian cycle of leptin is preserved in constitutional thinness whereas it is not in anorexia nervosa [14]. Similar to anorexia nervosa, individuals with constitutional thinness have lower bone mass (i.e., osteopenia) compared with controls [15,16]. Overall, most hormonal systems are unaffected by low body mass in constitutional thinness.

Human body weight regulation has a substantial genomic component [17] and the first genome-wide association study (GWAS) of constitutional thinness estimated its single nucleotide polymorphism-based heritability (SNP-based *h*^2^) at 28% [18], indicating that a substantial proportion of the trait variance is associated with common genomic variants. Constitutional thinness was defined as ≤ 18 kg/m^2^ and based on detailed medical and medicine history from general practioner records in British individuals aged 18-65 years. Individuals with chronic renal, liver, gastrointestinal, metabolic diseases, psychiatric disorders, or eating disorders, and high physical activity (i.e., >3 times per week) were excluded. Moreover, individuals were screened for eating disorders using the SCOFF questionnaire [19] and needed to endorse the question that they always had been thin to assess persistence. The study also revealed a genetic overlap with severe early-onset obesity, as the genetic correlation (*r*_g_) between the two traits was −0.49. This constitutional thinness phenotype showed a low statistically non-significant genetic correlation with anorexia nervosa (*r*_g_ = 0.13, *p* = 0.09) [18], potentially a power issue.

Anorexia nervosa shows genetic correlations with other psychiatric disorders, including obsessive-compulsive disorder, major depressive disorder, anxiety disorders, and schizophrenia [20–22]. Whether constitutional thinness shows a similar pattern of genetic correlations as seen in anorexia nervosa is unknown.

Our overall aim was to examine the differences between constitutional thinness and anorexia nervosa on a genomic level. For this, we used two genetically-informed approaches:

1. We hypothesised that constitutional thinness and anorexia nervosa would not be genetically correlated and that we would not see the same pattern of genetic correlations between constitutional thinness and psychiatric disorders that we see in anorexia nervosa.
2. Using polygenic scores, which aggregate summary statistics from previous GWASs into individual specific risk scores, we hypothesised that in our target population constitutional thinness would be associated with polygenic scores for constitutional thinness, obesity, and other anthropometry-related traits, but not with psychiatric disorders.

## Methods

### Linkage disequilibrium score regression to calculate genetic correlations

To estimate the genetic overlap between constitutional thinness/anorexia nervosa and psychiatric disorders, we obtained GWAS summary statistics from studies on constitutional thinness [18], attention deficit hyperactivity disorder [23], anorexia nervosa [20], anxiety disorders [24], autism spectrum disorder [25], bipolar disorder [26], borderline personality disorder [27], major depressive disorder [28], obsessive-compulsive disorder [29], posttraumatic stress disorder [30], alcohol dependence [31], and schizophrenia [32]. For a full list of the GWAS summary statistics, see **Supplementary Table S1**. We used linkage disequilibrium score regression, version 1.0, to calculate genetic correlations as previously described [33,34]. Pleiotropic genomic variants or correlated causal genomic loci can give rise to genetic correlations, meaning that two traits share genomic variants that can either influence a trait in the same direction (i.e., positive genetic correlation) or in opposite directions (i.e., negative genetic correlation) [35]. To estimate these genetic correlations, the effect size of the first trait at the corresponding genomic polymorphism is multiplied by the effect size of the second trait. The product is regressed on the linkage disequilibrium score and the slope reflects the genetic correlation between both traits [33,34].

### Sample from the Avon Longitudinal Study of Parents and Children (ALSPAC)

During the period April 1, 1991, until December 31, 1992, the ALSPAC cohort was started by inviting pregnant women in the former county of Avon, United Kingdom, to participate in the developmental population-based cohort [36–39]. The cohort originally included 14,541 pregnancies of whom 13,988 children were alive at one year and the cohort was boosted with 913 children at the age of seven years. For the exclusion of disordered-eating symptoms, follow-up was conducted at age 14 (wave 14, *n* = 10,581), 16 (wave 16, *n* = 9,702), and 18 years (wave 18, *n* = 9,505) with these response rates: 6,140 (58%) responding at wave 14, 5,069 (52%) at wave 16, and 3,228 (34%) at wave 18. Additionally, parent-reported questionnaires are available for 7,025 adolescents at wave 14 and on 5,656 at wave 16 to strengthen the validity of the probable eating disorder diagnoses [40]. Combining self-reports during adolescence and parent-reports, we assigned probable eating disorder diagnoses to adolescents [40], a gold standard for childhood psychiatric disorders (for diagnostic criteria, see **Supplementary Table S2**). Study data were collected and managed using REDCap electronic data capture tools hosted at University of Bristol [41,42]. REDCap (Research Electronic Data Capture) is a secure, web-based software platform designed to support data capture for research studies, providing 1) an intuitive interface for validated data capture; 2) audit trails for tracking data manipulation and export procedures; 3) automated export procedures for seamless data downloads to common statistical packages; and 4) procedures for data integration and interoperability with external sources. Please note that the study website contains details of all the data that is available through a fully searchable data dictionary and variable search tool (www.bristol.ac.uk/alspac/researchers/our-data/). To assure genetic unrelatedness of participants in our analyses, we selected randomly one individual of each pair that is closely related (*φ* > 0.2) using PLINK v1.90 [43], excluding 75 participants.

### Ethics approval and consent to participate

Ethical approval for the ALSPAC participants was obtained from the ALSPAC Ethics and Law Committee and the Local Research Ethics Committees: www.bristol.ac.uk/alspac/researchers/research-ethics/. The main caregiver initially provided consent for child participation, and from the age 16 years, the offspring themselves have provided informed written consent. At 16 years old, sole consent from the study child was considered acceptable by the Committee and although the law was not specific about young people with regards to research, this complied with the Family Law Reform Act 19695 as regards treatment: those who are 16 years old or above have the same legal ability to consent to any medical, surgical or dental treatment as anyone above 18, without consent from their parent or guardian [44]. Children were invited to give assent where appropriate. Consent for biological samples was collected in accordance with the Human Tissue Act (2004) and informed consent for the use of data collected via questionnaires and clinics was obtained from participants following the recommendations of the ALSPAC Ethics and Law Committee at the time. All methods were carried out in accordance with relevant guidelines and regulations.

### Genotyping, imputation and quality control in the Avon Longitudinal Study of Parents and Children (ALSPAC)

Genome-wide genotyping was performed on 9,915 of 15,247 children participating in ALSPAC using the Illumina HumanHap550 quad chip. Participants with a SNP missingness >3%, insufficient sample replication (identity by descent < 0.1), biological sex mismatch, and noneuropean ancestry (as defined by multi-dimensional scaling using the HapMap Phase II, release 22, reference populations) were excluded. All SNPs underwent the following quality control: They were excluded if their minor allele frequency (MAF) was < 1%, excessive missingness occurred (i.e., call rate < 95%), or a departure from the Hardy–Weinberg equilibrium had a *p* value < 5 × 10^−7^. Genotypes were imputed with Impute3 to the Human Reference Consortium (HRC) 1.0 reference panel [45] and phased using ShapeIT (v2.r644). After imputation, any SNPs with MAF < 1%, Impute3 information quality metric of < 0.8, and Hardy-Weinberg equilibrium *p* < 5 × 10^−7^ were excluded. After quality control, 8,654 participants (4,225 females and 4,429 males) and 4,054,653 SNPs could be carried forward for analyses.

### Latent class growth analysis to identify individuals with constitutional thinness in adolescence and young adulthood

We identified participants with constitutional thinness analysing objectively measured BMI (kg/m^2^) collected at 10, 12, 13, 14, 16, 18, and 24 years in ALSPAC [39]. Prior to analysis, BMIs at each time point were log-transformed and dichotomised in the bottom 5% versus remaining 95%. Participants who engaged in any weight loss behaviours (e.g., fasting, purging, and/or laxative use) at 14, 16, or 18 years, or met criteria for anorexia nervosa, bulimia nervosa, and/or purging disorder (*n* = 791) were excluded. For all participants who had at least one measure of BMI (*n* = 8,505), we derived longitudinal latent classes using latent class growth analysis with a full information maximum likelihood [46,47] approach on the log-transformed and dichotomised BMI categories in Mplus version 8.4. This approach allows the inclusion of all participants who had at least one data point, maximising our sample size.

The number of classes is not directly estimated and alternative specifications with increasing numbers of classes were compared. The best-fitting model was identified using the Akaike Information Criterion (AIC), Bayesian Information Criteria (BIC), and entropy. For AIC and BIC, lower values indicate better model fit, whereas a high entropy is desirable. Further, trajectory size is taken into consideration, as small (<3%) trajectories are hard to interpret. After selection of the optimal number of classes, estimations were repeated using 1000 random starts to avoid local maxima. A model with three classes was the best fit in comparison with the one, two, or four class solution (for a full list of fit statistics, see **Supplementary Table S3**). The majority (*n* = 7,884, 93%) of participants were never in the class of the lowest 5% of logBMI. Participants assigned to the second class (*n* = 349, 4%) had a 0.25 probability to be in the lowest 5% of the logBMI distribution, whereas participants in class three (*n* = 272, 3%) were highly likely to be in the lowest 5% of the logBMI distribution at all time points (**Figure 1**). Class three was considered to represent participants with constitutional thinness and classes one and two were collapsed creating the control group. The resulting two groups were carried forward in the following analyses: individuals with constitutional thinness (*n* = 272; 3%) and individuals without constitutional thinness (*n* = 8,233; 97%). For illustration, the mean BMI and standard deviation at each wave, for each class, are listed in **Supplementary Table S4**.

**Figure 1.**
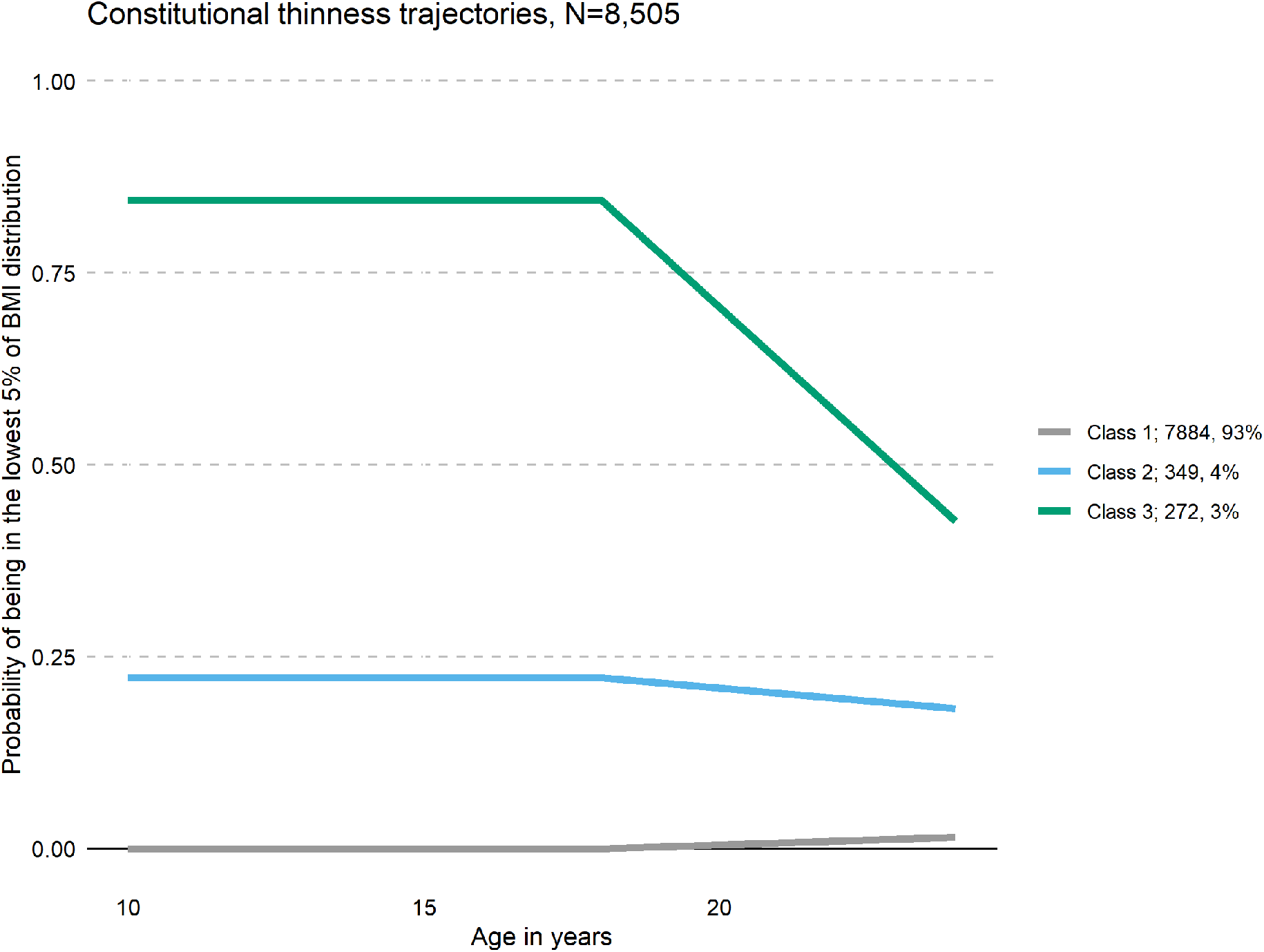
Results from the latent class growth analyses of constitutional thinness in the Avon Longitudinal Study of Parents and Children (ALSPAC, n = 8,505). Classes 1 and 2 were collapsed in our analysis as one comparison group (n = 8,156). We defined class 3 (green) as individuals with constitutional thinness in adolescence and in young adulthood in ALSPAC (n = 272).

### Polygenic risk scoring in ALSPAC

To calculate polygenic scores of psychiatric disorders and anthropometric traits, we used the software PRSice [48], version 2.2.3. During this procedure, to obtain genetically independent SNPs, SNPs present both in the discovery GWAS summary statistics and in the genotype data of the ALSPAC cohort (i.e., overlapping SNPs) were clumped. The SNP with the lowest *p* value in each 250 kilobase window of all those in linkage disequilibrium (*r*^*2*^ > 0.1) was carried forward. We calculated eleven polygenic scores for psychiatric disorders and 13 anthropometry-related polygenic scores, including BMI [49], overweight, obesity class 1-3, extreme BMI [50], hip and waist circumference [51], childhood obesity [52], age at menarche [53], and constitutional thinness [18] at their optimal *p* value threshold in each individual of our subsample (for a full list of discovery GWAS, see **Supplementary Table S1**). For this, we weighted the number of effect alleles by the corresponding allele effect size across the remaining SNPs. We used high-resolution scoring (i.e., incrementally across a large number of *p* value thresholds) to obtain the *p* value threshold at which the polygenic score is optimally associated with the outcome and explains the most variance (i.e., resulting in Nagelkerke’s *R*^*2*^ on the liability scale). We fitted logistic regressions with constitutional thinness as the outcome, adjusted for sex and the first six principal components. We calculated empirical *p* values by permuting case-control status at every *p* value threshold 10,000 times to account for potential overfitting. We converted the observed *R*^*2*^ to the liability scale using the sample prevalence of constitutional thinness as population prevalence.

We calculated *Q* values using the false discovery rate approach [54,55] to correct for 24 polygenic score regression models. We did not stratify analyses by sex due to the few individuals with constitutional thinness but included sex as a covariate.

## Results

### Genetic correlation between constitutional thinness and anorexia nervosa

The genetic correlation between the anorexia nervosa GWAS [20] and the constitutional thinness GWAS [18] was modest and not statistically significant (*r*_g_ = 0.13, se = 0.08, *p* = 0.09).

### Genetic correlations between constitutional thinness and other psychiatric disorders

In contrast to anorexia nervosa, constitutional thinness showed a statistically significant negative genetic correlation with attention deficit hyperactivity disorder (*r*_g_ = −0.24, se = 0.09, *Q* = 0.04) and a much stronger negative correlation with alcohol dependence (*r*_g_ = −0.44, se = 0.16, *Q* = 0.04, **Figure 2**). Anorexia nervosa, in turn, showed positive genetic correlations with lifetime anxiety disorders (*r*_g_ = 0.25, se = 0.05, *Q* = 6.2 × 10^−7^), autism spectrum disorder (*r*_g_ = 0.20, se = 0.04, *Q* = 2.7 × 10^−4^), bipolar disorder (*r*_g_ = 0.18, se = 0.04, *Q* = 4.8 × 10^−6^), borderline personality disorder (*r*_g_ = 0.23, se = 0.09, *Q* = 0.03), major depressive disorder (*r*_g_ = 0.28, se = 0.07, *Q* = 3.1 × 10^−4^), obsessive compulsive disorder (*r*_g_ = 0.45, se = 0.08, *Q* = 5.2 × 10^−8^) and schizophrenia (*r*_g_ = −0.24, se = 0.09, *Q* = 9.6 × 10^−17^). In summary, constitutional thinness shows negative genetic correlations with one cluster of psychiatric disorders while anorexia nervosa shows positive genetic correlation with another cluster. For all calculated genetic correlations, see **Supplementary Table S5**.

**Figure 2.**
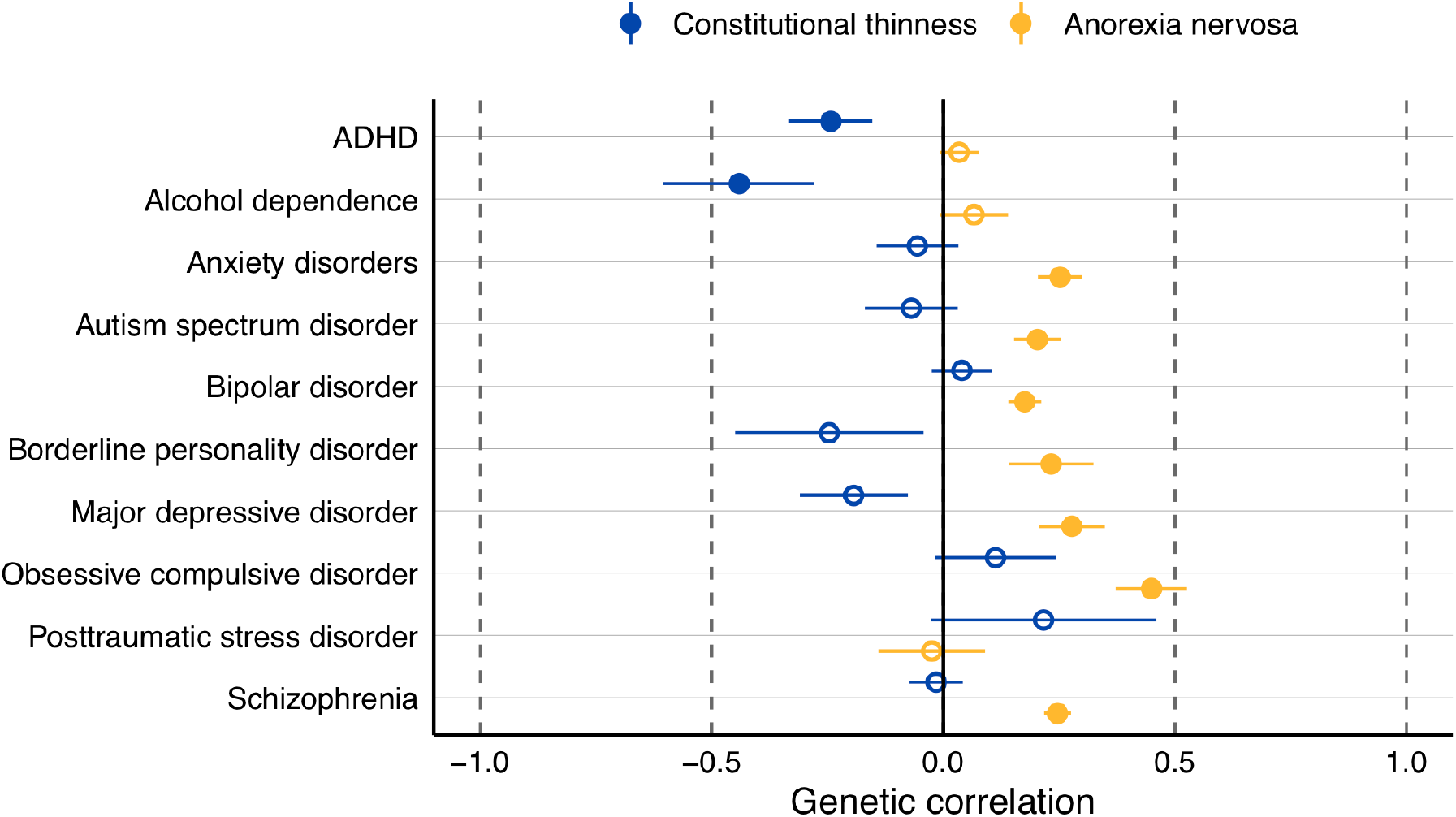
Genetic correlations of constitutional thinness and anorexia nervosa. The plot shows genetic correlations of constitutional thinness (blue) and anorexia nervosa (yellow) with psychiatric disorders calculated by linkage disequilibrium score regression. Filled dots are statistically significant after adjustment for multiple testing through the false discovery discovery approach. Dots represent genetic correlations (r_g_) and error bars index standard errors. ADHD = attention deficit hyperactivity disorder

### Polygenic scores associated with constitutional thinness in adolescence and young adulthood

The effect sizes are expressed as odds ratios (OR) per standard deviation increase of polygenic score (for a full list of results, see **Supplementary Table S6**). It is important to note that there was no evidence of an association between constitutional thinness polygenic score (OR = 1.16, 95% CI: 1.00, 1.33; *Q* = 0.48) and constitutional thinness in adolescence and young adulthood in ALSPAC.

An increased genetic predisposition for borderline personality disorder (OR = 0.77, 95% CI: 0.67; 0.88; *Q* = 0.01) was associated with lower risk of constitutional thinness during adolescence and young adulthood in ALSPAC (**Figure 3, Panel A**), whereas a higher genetic predisposition for posttraumatic stress disorder (OR = 1.27, 95% CI: 1.11; 1.46; *Q* = 0.01) was associated with a higher risk of thinness.

**Figure 3.**
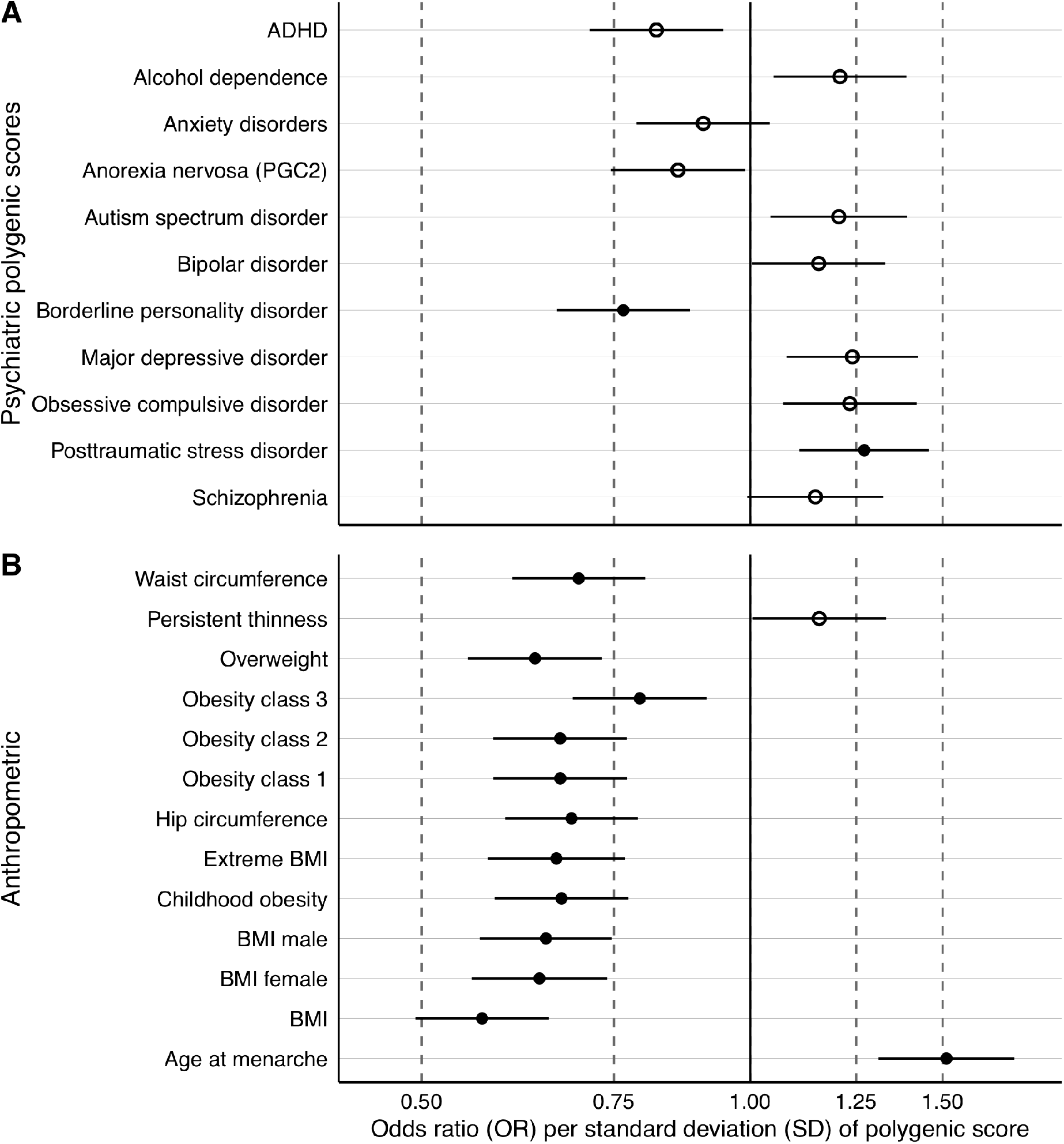
**Panel A & B** Polygenic scores associated with constitutional thinness in the Avon Study of Parents and Children (ALSPAC). **Panel A** shows psychiatric and **Panel B** anthropometric polygenic scores and their association estimates with latent class growth analysis-derived constitutional thinness (n = 8,505). Filled dots are statistically significant after adjustment for multiple testing through the false discovery discovery approach. Dots represent odds ratios (ORs) and error bars index 95% confidence intervals obtained via logistic regression and 10,000 permutations to obtain empirical p values. ADHD = attention deficit hyperactivity disorder, BMI = body mass index, PGC2 = second freeze of the anorexia nervosa genome-wide association study of the Psychiatric Genomics Consortium

Nine anthropometric polygenic scores were negatively associated with constitutional thinness in adolescence and young adulthood (i.e., age 10-24 years; **Figure 3, Panel B** and see **Supplementary Table S6** for full results). A genetic predisposition to a higher BMI (OR = 0.57, 95% CI: 0.49, 0.65; *Q* = 2.4 × 10^−4^) was associated with a lower risk of constitutional thinness in adolescence and young adulthood. Overall, the associations with anthropometric polygenic scores were stronger than with psychiatric polygenic scores. Consistently, a higher genetic predisposition to childhood obesity (OR = 0.67, 95% CI: 0.58; 0.77; *Q* = 2.4 × 10^−4^) was associated with a lower risk of constitutional thinness in adolescence and young adulthood. The childhood (OR = 0.67) and adulthood (OR = 0.67) obesity polygenic scores showed similar effect sizes of their association with constitutional thinness with overlapping confidence intervals (CIs). Similarly, BMI polygenic scores based on men or women only showed no differences in the effect sizes of the associations (OR_men_ = 0.65; OR_women_ = 0.64 with overlapping CI), suggesting no differences between sexes. Additionally, the age at menarche polygenic score was positively associated with constitutional thinness (OR = 1.51, 95% CI: 1.31, 1.74; *Q* = 2.4 × 10^−4^) in contrast to the other anthropometric polygenic scores.

## Discussion

In the first part of our study and in line with our hypotheses and previous research [18], we demonstrated that constitutional thinness shows no statistically significant genetic correlation with anorexia nervosa, indicating that they are genomically distinct conditions. Contrary to our hypothesis, however, two of eleven psychiatric disorders, attention deficit hyperactivity disorder and alcohol dependence, showed a negative genetic correlation with constitutional thinness. This means that attention deficit hyperactivity disorder and alcohol dependence share genetic variants with constitutional thinness that influence both traits in opposite directions.

In the second part of our study using polygenic scores, we showed that individuals who were persistently thin from age of 10 years until 24 years (defined as being longitudinally more likely to be in the lowest 5% of the logBMI distribution) carried lower polygenic scores for borderline personality disorder, but higher polygenic scores for posttraumatic stress disorder than individuals in the remaining 95% of the logBMI distribution. This means that against our hypothesis, individuals with constitutional thinness during adolescence and young adulthood may carry genomic variants that predispose them to the development of a posttraumatic stress disorder.

These persistently thin individuals also carried lower polygenic scores for anthropometry-related polygenic scores, including BMI, hip, and waist circumference, and higher age at menarche polygenic scores than individuals with higher BMIs. This is expected because BMI and age at menarche are negatively genetically correlated [56]. Most importantly, we did not observe an association between the polygenic score for constitutional thinness and our derived constitutional thinness phenotype in the ALSPAC cohort. We hypothesise that this may be either due to the age mismatch between the discovery GWAS (i.e., 18-64 years) and our constitutional thinness phenotype (i.e., 10-24 years), as GWASs of BMI are not perfectly correlated across age [22] or due to limited statistical power to detect an association as both ALSPAC (*n* = 272) and the discovery GWAS (*n* = 1471) comprised relatively few individuals with constitutional thinness.

We did not observe any differences in anthropometric polygenic scores when based on female only or male only GWAS of BMI or when the polygenic scores were derived from childhood or adulthood obesity GWASs, suggesting no sex or age differences in the genetic associations of anthropometry-related polygenic scores with constitutional thinness.

Phenotypically, individuals with constitutional thinness do not demonstrate psychopathology or disordered-eating behaviours [3]. Our analysis showed genetic evidence that constitutional thinness is associated with lower risk for attention deficit hyperactivity disorder, alcohol dependence, and borderline personality disorder. All three psychiatric disorders are characterised by impairments in inhibitory control [57–61]. We hypothesise that the negative genetic associations may reflect genetic underpinnings for elevated inhibitory control potentially explaining the lower calorie intake observed in constitutional thinness [4]. We hypothesise that this shared phenotypic characteristic between constitutional thinness and the disinhibition-related psychiatric disorders may explain the negative genetic overlap. However, GWAS investigating inhibitory control are necessary to investigate this overlap directly on a genomic level and answer the question of which trait or traits may be responsible for the shared genetics between disinhibition-related psychiatric disorders and constitutional thinness.

Previous research revealed a negative genetic correlation between constitutional thinness and BMI [18]. We replicated this finding in our polygenic score analysis of constitutional thinness in adolescence and young adulthood in ALSPAC. It is important to note that anorexia nervosa also shows a negative genetic correlation with BMI [20,62]. However, constitutional thinness and anorexia nervosa are not directly genetically correlated in our analysis and previously [18], suggesting that two different sets of genetic variants may be implicated in the shared genetics between constitutional thinness and BMI, and between anorexia nervosa and BMI. Alternatively, both the estimation of the genetic correlation as well as the polygenic score association may be underpowered as the number of individuals with constitutional thinness is small in the discovery GWAS and the ALSPAC cohort.

Another aspect in which constitutional thinness differs from anorexia nervosa is its negative genetic overlap with attention deficit hyperactivity disorder. Anorexia nervosa has not been reported to correlate genetically with attention deficit hyperactivity disorder [20,63]. However, attention deficit hyperactivity disorder and constitutional thinness are genetically correlated with BMI [23,62]. This suggests that a set of genetic variants exists that may explain the shared genetics among constitutional thinness, BMI, and attention deficit hyperactivity disorder that is distinct from the shared genetics between anorexia nervosa and BMI.

In contrast to our study, many genetic studies have focused on high weight and obesity with considerably less attention paid to the lower end of the weight spectrum. Our analysis benefitted from repeated measurements across development that afforded a more robust phenotype than cross-sectional thinness measures. Furthermore, we reduced the probability of reporting false positives because we excluded individuals with disordered-eating behaviour and eating disorders from our polygenic score analyses.

Limitations regarding our constitutional thinness phenotype definition in ALSPAC must be acknowledged. Some participants may only develop obesity later in life [17,64] and, as our longitudinal modelling ended at age 24 years, some individuals may not continue to remain thin. Individuals taking medication or with physical conditions that affect weight regulation, or who are using non-prescription illicit substances, or smoke [17] were not excluded. In addition, by nature of the available data, we only investigated genomics in white British participants. It is essential that future studies include large diverse populations to ensure that our genetic investigations accurately capture the ways in which genes influence disease risk in ancestrally diverse populations.

Confirming our hypothesis, constitutional thinness (characterized by persistent low BMI and no disordered-eating behaviors or cognitions) was significantly negatively associated with anthropometric polygenic scores. Additionally, we found that constitutional thinness was negatively genetically associated with psychiatric disorders characterised by impaired inhibitory control, such as attention deficit hyperactivity disorder, alcohol dependence, and borderline personality disorder, suggesting that individuals with constitutional thinness may carry a lower number of genetic risk variants for those disorders. However, our observations clearly delineate constitutional thinness from anorexia nervosa, which shows a different pattern of positive genetic correlations with psychiatric disorders (i.e., with obsessive-compulsive disorder, schizophrenia, major depressive disorder, anxiety disorders, and bipolar disorder) [20,26,34]. Our findings confirm that constitutional thinness and anorexia nervosa are largely separate entities that may be characterized by differential underlying genomic liability for psychiatric illness and body weight regulation.

## Data Availability

This study is based on data from the ALSPAC study (http://www.bristol.ac.uk/alspac/). Interested researchers can apply for data access with the University of Bristol, UK. Analysis scripts can be requested from the authors.

http://www.bristol.ac.uk/alspac/

## Acknowledgements

We are extremely grateful to all the families who took part in this study, the midwives for their help in recruiting them, and the whole ALSPAC team, which includes interviewers, computer and laboratory technicians, clerical workers, research scientists, volunteers, managers, receptionists and nurses. This study was completed as part of approved UK Biobank study applications 27546 to Prof Breen.

## Author Contribution

CH, MA, MH, ABP analysed the data. CH, MA, MH drafted the manuscript. CMB, NM, RJFL, and GB supervised the work. All authors substantially contributed to the conception and interpretation of the work, revised the manuscript for important intellectual content and approved the final version. All authors agree to be accountable for all aspects of this work.

